# Frequent, Dual and Nighttime Nicotine Use Among Dutch Adolescents: Findings From a School-Based Survey

**DOI:** 10.1101/2025.11.26.25341067

**Authors:** Daphne C.J. Raad, Anne Marit Koome, Yousef el Baser, Frank J. Borm, Esther A. Croes, Danielle Cohen, David van Bodegom

## Abstract

**Purpose:** Adolescent nicotine use has increased recently. Detailed insights on usage patterns remain limited but are essential for effective prevention and intervention strategies. This study provides recent school-based data to enhance understanding of nicotine use among Dutch secondary school students.

**Methods:** A cross-sectional survey was conducted among students from five Dutch secondary schools between March and May 2025. Data were collected using a self-report questionnaire administered both as a school-wide survey and during sessions with students specifically identified as nicotine users. Measures included demographics, nicotine use patterns (age at initiation, product type, frequency), perceived parental awareness, perceived addiction, quit attempts, and self-reported health complaints.

**Results:** Of 978 respondents (mean age 14.5 years), 457 reported ever using nicotine, with 90% of these reporting past-year use. Median initiation age was 13 years. E-cigarettes were the predominant initial product (80%), followed by tobacco cigarettes (18%) and nicotine pouches (2%). Past year use was dominated by e-cigarettes (83%) and tobacco cigarettes (74%). Nearly half (47%) of past-year users reported daily use, over half (53%) used nicotine during school hours, and over one-third (35%) used nicotine upon waking at night. Among these past-year users, 32% perceived parental unawareness, 41% perceived themselves as addicted, 60% reported quit attempts, most unsuccessful, and 61% reported health complaints attributed to nicotine use.

**Conclusion:** Adolescent nicotine use is characterized by frequent daily use, including use during school hours and nighttime awakenings, limited parental awareness, perceived addiction, largely unsuccessful quit attempts, and notable health complaints. These findings emphasize the urgent need for school-based cessation programmes involving parents, combined with policies to limit tobacco industry influence and reduce youth nicotine access.

**WHAT IS KNOWN:**

- Adolescent nicotine use has increased recently.
- National surveys provide prevalence estimates but lack detailed data on usage patterns.

**WHAT IS NEW:**

- Frequent daily use (47%) and nicotine use during nighttime awakenings (35%), perceived addiction (41%), and quit attempts (60%), with high rates of self-reported health complaints (61%), indicate substantial nicotine dependence.
- E-cigarettes are the predominant initiation product (80%) compared to cigarettes (18%), while past-year use involves both e-cigarettes (83%) and cigarettes (74%), supporting the gateway effect.
- High school-hour use and limited perceived parental awareness stress the need for school-based cessation interventions.
- Most adolescents begin use around age 13, often progressing to regular use, underscoring policymakers’ responsibility to limit the tobacco industry’s influence and reduce youth access to nicotine products.

## INTRODUCTION

Nicotine use among adolescents has increased in recent years, largely driven by the widespread availability of electronic cigarettes (e-cigarettes) (1-3). Although these products were initially marketed as smoking cessation aids for adult smokers, they are frequently flavoured and promoted in ways that appeal to young people, such as through social media influencers (4-7). Rather than serving as effective smoking cessation tools, e-cigarettes may act as a gateway to nicotine initiation among youth, fostering dual use of e-cigarettes and tobacco cigarettes (8-13). Recent longitudinal data from the United Kingdom indicate that the predicted probability of cigarette smoking ranges from 1% among e-cigarette naïve youth to 33% among current e-cigarette users (14). Also, a high rate of dual use has been observed in the Netherlands, where 69% of monthly e-cigarette users also report monthly cigarette smoking (15). Furthermore, the assumption that e-cigarettes are a harmless alternative to tobacco cigarettes is increasingly challenged by growing evidence of their health risks. Both products deliver nicotine, a highly addictive substance associated with significant adverse health effects, particularly in youth, including increased heart rate, respiratory symptoms, impaired impulse control, and negative effects on mental health (7, 16-19). While the long-term harms of cigarette smoking are well established and severe, emerging evidence suggests that e-cigarettes, through acute toxicity and the inhalation of harmful substances such as heavy metals and carcinogens, may also increase the risk of chronic cardiovascular and respiratory diseases, cancers, depression, anxiety, and potentially impair brain development during adolescence (7, 16, 19-24). These serious health concerns underscore the need for comprehensive data on adolescent nicotine use.

In the Netherlands, effective monitoring of adolescent nicotine use has a longstanding history. Since 1984, the Trimbos Institute has collected nationally representative data on substance use (tobacco, alcohol, and drugs), social media use, gaming, gambling, mental health, and school experience among 12- to 16-year-olds. The most recent data available show that in 2023, 25% reported ever using e-cigarettes and 16% ever smoking cigarettes, with 4% and 2% respectively reporting daily use. Additionally, 4% of this age group reported ever using nicotine pouches (2, 15). However, detailed data on nicotine usage patterns remain lacking, yet are crucial for improving understanding of adolescent nicotine behaviour and for guiding effective prevention and cessation strategies.

This study aims to provide detailed data on nicotine use patterns, perceived parental awareness, and self-reported health complaints among Dutch secondary school students. By surveying nearly 1,000 students across five secondary schools, this study offers comprehensive insights to better inform health professionals and policymakers in developing targeted prevention and cessation strategies to address this growing public health concern.

## METHODS

### Study Design

This cross-sectional survey was conducted between March and May 2025 across five Dutch secondary schools that voluntarily participated in a broader pilot project of Vaping #YourChoice (Dutch: Vapen #JouwKeuze), a Dutch foundation providing free educational packages to schools aimed at preventing adolescent nicotine use (25). The socioeconomic context of each school was quantified using the Educational Disadvantage Index (Dutch: Achterstandsscore), developed by Statistics Netherlands (CBS). This index reflects the risk of educational disadvantage within school populations, with scores ranging from 0 to 981, where higher scores indicate greater disadvantage.

### Data Collection

The target study population consisted of all students enrolled at the participating schools during the data collection period. Data collection took place during the period when final-year students were occupied with their exam schedules, which limited their availability and accessibility. Data were collected using two complementary recruitment strategies via self-report questionnaires: a school-wide survey and a targeted survey administered during informational sessions with students identified as nicotine users.

1. School-wide surveys: Online or paper-based questionnaires were distributed to all students by school staff.
2. Targeted surveys: Paper-based questionnaires were administered during informational sessions initiated by *Vapen #jouwkeuze*. Students identified by school staff as active nicotine users (e.g., those observed using nicotine during school breaks) were invited to attend these sessions.

### Measures

Survey items were adapted from the Trimbos Institute’s Youth Tobacco and Nicotine Products Monitor (15). The surveys contained 22 items (see **Appendix 1**). Educational tracks were classified following Trimbos Institute practices, grouping combined tracks under the lower-level category (e.g., HAVO/VWO classified as HAVO) (15). Consequently, the main categories were VMBO, HAVO, and VWO. Key measures included:

- Demographics: age, sex, school, grade, and educational track
- Nicotine use: lifetime and past-year use
- Usage patterns: age at initiation, product type, frequency of use (overall, during school hours, and during nighttime awakenings), and perceived parental awareness
- Perceived health problems: self-reported health complaints attributed to nicotine use, perceived addiction, quit attempts (both successful and unsuccessful), and cessation intentions

### Ethical Considerations

Participation in the study was voluntary and anonymous. Students were provided with clear information regarding the study’s purpose, data confidentiality, and their right to withdraw at any time without consequence. Parents were informed through official school communication channels. The Medical Ethical Review Board of Leiden University Medical Centre confirmed that this research does not fall under the Dutch Medical Research with Human Subjects Act (WMO), as it does infringe upon the physical or psychological integrity of participants. Consequently, the non-WMO Review Committee reviewed the study protocol and provided a declaration of no objection (reference number 25-3070). The study was conducted in accordance with the Declaration of Helsinki (2008 revision). All procedures adhered to ethical standards appropriate for a non-clinical, voluntary, observational study.

### Data Analysis

Data were analysed descriptively using R (version 4.4.1; R Core Team, Vienna, Austria, 2025) to summarise frequencies, proportions, and nicotine use patterns. Analyses were stratified by recruitment method (school-wide versus targeted surveys). Percentages were calculated based on the number of valid responses per question, excluding missing data. Given the survey design, only participants reporting lifetime nicotine use were asked about past-year use. Hence, past-year use prevalence is presented as a proportion of the entire study sample rather than only among responders to that item. Participants who completed only the initial screening questions without further responses were excluded to maintain data completeness and validity.

## RESULTS

### Response rates

A total of 978 student questionnaires were collected from five Dutch secondary schools, representing a combined student population of approximately 3,550. Response rates, calculated as the proportion of responses relative to the total student population per school, varied by school and ranged from 22% to 39%. The largest school (School 2), with around 2,000 students, contributed 437 responses across both survey modes, whereas the smallest school (School 5), a school specialised in education for students with severe intellectual or multiple disabilities and comprising approximately 50 students, provided 14 responses. The participating schools represented a range of socioeconomic contexts, generally reflecting relatively favourable conditions, with Achterstandsscores ranging from 1 to 128. All of the schools offered the full spectrum of Dutch secondary education tracks, VMBO (pre-vocational secondary education), HAVO (senior general secondary education), and VWO (pre-university education). Further details on participation rates and school characteristics are presented in **Table 1**.

**Table 1.** Responses per school and recruitment strategy.

|  | School 1 | School 2 | School 3 | School 4 | School 5 |
| --- | --- | --- | --- | --- | --- |
| Number of students (approx.) | 700 | 2000 | 200 | 600 | 50 |
| Educational tracks | VMBO | VMBO | VMBO | VMBO | VMBO |
|  | HAVO | HAVO | HAVO | HAVO | HAVO |
|  | VWO | VWO | VWO | VWO | VWO |
| Educational Disadvantage Index<sup>1</sup> | 104 | 128 | 1 | 23 | Unavailable |
| Targeted survey responses | 64 | 93 | 27 | 73 | 4 |
| Of whom using nicotine<sup>2</sup> | 54 | 78 | 17 | 45 | 4 |
| School-wide survey responses | 164 | 344 | 37 | 162 | 10 |
| Of whom using nicotine<sup>2</sup> | 64 | 76 | 11 | 40 | 7 |

<sup>1</sup> Educational Disadvantage Index (Dutch: Achterstandsscore): an indicator developed by Statistics Netherlands (CBS) reflecting the risk of educational disadvantage within a school population based on student background factors such as parental education, home language, and length of residence in the Netherlands. Scores range from 0 to 981, with higher values indicating greater risk.
<sup>2</sup> Answered “yes” on self-reported nicotine use in the past 12 months.
Abbreviations: VMBO, pre-vocational secondary education; HAVO, senior general secondary education; VWO, pre-university education.

### Participant characteristics

The school-wide surveys yielded 717 responses. Among these respondents, 251 students reported ever using nicotine and 198 students (28% of the school-wide sample) reported nicotine use within the past 12 months. The mean age of students in the school-wide sample was 14.3 years (SD = 1.5). Approximately 54% identified as female, 41% as male, and the remainder as ‘other’ or preferred not to disclose. Grade-level distribution included 19% in grade 1, 22% in grade 2, 39% in grade 3, 13% in grade 4, 5% in grade 5, and 2% in grade 6. Educational tracks included VMBO (29%), HAVO (41%), and VWO (31%).

The targeted surveys yielded 261 responses. Among these respondents, 206 students reported ever using nicotine and 198 students (76% of the targeted sample) reported nicotine use within the past 12 months. The mean age of students in the targeted sample was 14.9 years (SD = 1.2). Approximately 52% identified as female, 44% as male, and the remainder as ‘other’ or preferred not to disclose. Grade-level distribution included 7% in grade 1, 28% in grade 2, 33% in grade 3, 25% in grade 4, less than 1% in grade 5, and 7% in grade 6. Educational tracks included VMBO (47%), HAVO (35%), and VWO (18%). Characteristics of our two study samples are shown in **Table 2**.

**Table 2.** Characteristics of students by recruitment strategy.

| Characteristic of students | Total<br>(n = 978) | Students from school-wide survey (n = 717) | Students from targeted<br>survey (n = 261) |
| --- | --- | --- | --- |
| Mean age; years (SD) | 14.5 (1.4) | 14.3 (1.5) | 14.9 (1.2) |
| Sex |  |  |  |
| Female | 53% | 54% | 52% |
| Male | 42% | 41% | 44% |
| Other | 3% | 4% | 2% |
| Prefer not to say | 2% | 2% | 3% |
| Grade |  |  |  |
| 1 | 19% | 19% | 7% |
| 2 | 22% | 22% | 28% |
| 3 | 39% | 39% | 33% |
| 4 | 13% | 13% | 25% |
| 5 | 5% | 5% | 0% |
| 6 | 2% | 2% | 7% |
| Educational track |  |  |  |
| VMBO | 34% | 29% | 47% |
| HAVO | 39% | 41% | 35% |
| VWO | 27% | 31% | 18% |
| Ever used nicotine | 457 (47%) | 251 (35%) | 206 (79%) |
| Cigarettes (ever) | 74% | 70% | 80% |
| E-cigarettes (ever) | 89% | 85% | 93% |
| Nicotine pouches (ever) | 30% | 28% | 34% |
| Unknown (ever) | 5% | 6% | 3% |
| Median age first use; years (IQR) | 13 (2) | 13 (2) | 13 (1) |
| Cigarettes (first)* | 18% | 22% | 13% |
| E-cigarettes (first)* | 80% | 76% | 85% |
| Nicotine pouches (first)* | 2% | 2% | 2% |
| Past-year nicotine use | 396 (41%**) | 198 (28%**) | 198 (76%**) |
| Cigarettes (past-year) | 74% | 66% | 82% |
| E-cigarettes (past-year) | 83% | 77% | 89% |
| Nicotine pouches (past-year) | 25% | 23% | 27% |
| Unknown (past-year) | 7% | 11% | 3% |

\* Only students who indicated exactly one product are included. Multiple product (n = 102) and missing responses (n = 14) are excluded from percentages.
\*\* Percentages are calculated based on the number of respondents per category, except for past-year nicotine use. Past-year use was only asked of students who responded “yes” to ever using nicotine; therefore, these percentages are presented as proportions of total respondents.
Abbreviations: HAVO, senior general secondary education; IQR, interquartile range; SD, standard deviation; VMBO, pre-vocational secondary education; VWO, pre-university education.

### Using patterns of nicotine using students

Across both recruitment strategies, 457 students reported ever using nicotine, with a mean age of 14.9 years (SD = 1.5). Of these, 396 (90%) reported nicotine use within the past 12 months. The median age at initiation was 13 years (interquartile range (IQR) = 2), with e-cigarettes being the predominant initial product (80%), followed by cigarettes (18%) and nicotine pouches (2%). Past year use was characterised by multiple forms of nicotine use, with the majority reporting e-cigarette use (83%) and cigarette smoking (74%), and one-quarter (25%) reporting nicotine pouch use (**Table 2**).

Among past-year users, almost half (47%) reported daily use, 20% weekly use, and 9% monthly use, with the remainder reporting less frequent use or uncertainty regarding frequency (**Table 3**). More than half (53%) reported nicotine use during school hours, about one-quarter (24%) reported occasional use at school, and the remainder rarely or never used nicotine during school time. Nighttime nicotine use upon awakening was reported by more than one-third (35%) of users, with 26% affirming “yes” and 9% “sometimes,” while 65% rarely or never engaged in such use. Regarding perceived parental awareness, 44% of past-year users stated their parents were aware of their nicotine use, 32% perceived their parents as unaware, and the remainder were uncertain.

**Table 3.**
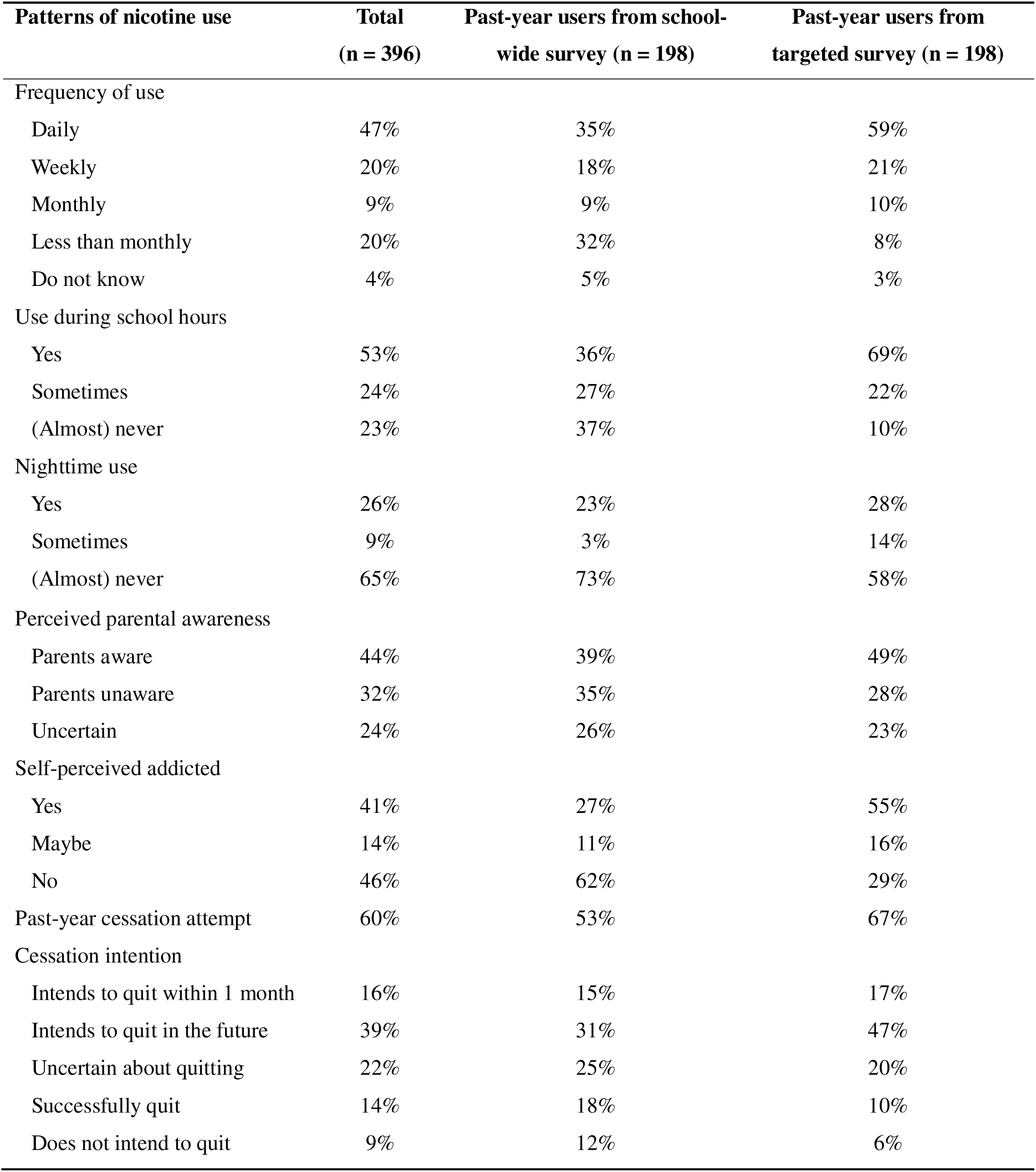
Patterns of nicotine use of past-year users by recruitment strategy.

Among past-year users, 41% self-identified as addicted by responding “yes” to the question asking whether they consider themselves addicted, 46% did not perceive themselves as addicted, and 14% were uncertain. Over half (60%) attempted to quit in the past year, with the majority of attempts unsuccessful. Of all past-year users, 14% reported successfully quitting, 9% expressed no intention to quit, 22% were uncertain about quitting, 39% planned to quit in the future, and 16% intended to quit within the next month (**Table 3**).

### Self-reported health complaints

A substantial proportion (61%) reported health complaints which they personally attributed to their nicotine use. The most frequently reported problems included reduced physical fitness (43%), coughing (32%), and sore throat (30%), followed by concentration difficulties (21%), headache (20%), nausea (19%), and sleep disturbances (16%). Smaller proportions reported increased depressive symptoms (12%) and anxiety symptoms or panic attacks (10%), with exacerbations of asthma (5%) and pneumonia (4%) being less common (**Table 4**).

**Table 4.** Self-reported health complaints among past-year nicotine-users (n = 365)

| Health complaints | Number of respondents |
| --- | --- |
| Any health complaint | 224 (61%) |
| Reduced physical fitness | 158 (43%) |
| Cough | 118 (32%) |
| Sore throat | 109 (30%) |
| Concentration difficulties | 78 (21%) |
| Headache | 74 (20%) |
| Nausea | 69 (19%) |
| Sleep disturbances | 59 (16%) |
| Increased depressive symptoms | 42 (12%) |
| Increased anxiety symptoms or panic attacks | 38 (10%) |
| Asthma exacerbation | 17 (5%) |
| Pneumonia | 13 (4%) |
| Unspecified | 3 (1%) |

## DISCUSSION

This study, with data from 2025, reveals frequent nicotine use among Dutch secondary school students, with 47% of users reporting daily consumption, 77% using nicotine during school hours, and 35% upon waking at night. Perceived parental awareness of nicotine use was limited to 44%. A substantial proportion (41%) perceived themselves as addicted, and 60% had attempted to quit in the past year, most without success. More than half (61%) reported health complaints that they attributed to nicotine use. Among users (mean age 14.9 years), initiation typically occurred around age 13, and 90% of ever-users reported past-year use, indicating that experimentation often progresses to regular use. The initial product used was predominantly e-cigarettes (80%) rather than tobacco cigarettes (18%), while past-year nicotine use was primarily with both e-cigarettes (83%) and tobacco cigarettes (74%). Collectively, these findings demonstrate substantial nicotine dependence and highlight multiple concerning patterns of adolescent nicotine use.

### Nicotine use during nighttime awakenings

During school-based consultations with adolescents as part of the Dutch *Vapen #jouwkeuze* prevention programme, where several authors (DC, FB, DR, YB, AK) are involved and physicians provide nicotine education (26), many students reported waking at nighttime due to nicotine cravings. This behaviour may indicate a more intense development of dependence associated with e-cigarette use. To explore this phenomenon, a specific item assessing nighttime nicotine use was incorporated into the survey. Nighttime use could function as a novel indicator of nicotine dependence in this age group, as it may reflect both physiological nicotine cravings and psychological distress (27).

The finding that over one-third of respondents (35%) reported using nicotine upon nighttime awakening, with 26% answering “yes” and 9% “sometimes”, is notable. The remaining 65% reported rarely or never engaging in such use. These unexpectedly high rates raise significant concerns regarding the severity of dependence. Also, our data reflected the substantial challenges adolescents face in seeking parental support or discussing their nicotine dependence. These findings underscore the urgent need for adults, including parents, educators, and healthcare providers, to take a proactive role in supporting youth struggling with nicotine addiction.

### Health implications of nicotine dependence

Beyond nighttime use, our findings on nicotine use patterns among current users suggest a considerable risk of dependence, indicated by frequent daily consumption, self-perceived addiction, and numerous unsuccessful quit attempts. Nicotine dependence harmfully affects both physical and mental health. Consistent with existing literature on the acute and chronic harms of nicotine products (7), a considerable proportion (61%) of adolescents in our study reported health complaints they attributed to nicotine use, including reduced physical fitness, respiratory symptoms, and cognitive difficulties.

Recent media reports in the Netherlands have highlighted severe cases involving adolescents hospitalized with pneumothorax, pulmonary haemorrhage, and intensive care unit (ICU) admissions related to e-cigarette use (28). These serious events likely represent the tip of the iceberg. Our study reveals that beneath these acute cases, many adolescents experience subclinical but significant health impairments. For instance, reduced physical fitness may indicate early alterations in lung function or impaired respiratory capacity that do not yet affect daily activities but become apparent during physical exertion. Given the current uncertainties about the exact causes of severe e-cigarette or vaping product use-associated lung injuries (EVALI), any adolescent presenting with e-cigarette-associated symptoms should be considered at risk (29). Our findings, which highlight the prevalent adverse effects of nicotine use on physical health alongside the serious health risks involved, underscore the need for increased clinical awareness and targeted preventive measures addressing adolescent nicotine use.

### Gateway hypothesis

In the Netherlands, nicotine products are prohibited for minors, and flavoured e-cigarettes have been banned for all ages since July 2024. Despite these regulations, adolescents continue to access these products through alternative channels, including social media platforms such as Snapchat and illicit sales via retail outlets (30). Our findings reinforce concerns regarding the role of e-cigarettes as a gateway to tobacco cigarette smoking (8-13). In our study, adolescents were specifically asked which nicotine product they first experimented with, with the majority (80%) reporting e-cigarettes as their initial product, while only 18% reported tobacco cigarettes. However, tobacco cigarettes were more frequently reported as used in the past year, primarily alongside e-cigarettes (74% and 83%, respectively). This aligns with longitudinal data from the United Kingdom, which indicate that the probability of cigarette smoking increases from 1% among e-cigarette naïve youth to 33% among current e-cigarette users (14). Our data provide further support for the gateway hypothesis within the Dutch adolescent population.

### Contexts for intervention

Despite frequent quit attempts, achieving sustained nicotine abstinence remains challenging among the adolescents surveyed. More than half (55%) expressed an intention to quit at the time of data collection, underscoring the need for cessation support tailored specifically to this age group. The high prevalence of nicotine use during school hours points to schools as critical intervention settings. Furthermore, limited perceived parental awareness, reported as absent or uncertain by over half of users (56%), suggests that family engagement should be an integral component of prevention and cessation programs, as parental engagement might play an important role in addressing adolescent substance use (31).

### Strengths and limitations

This study complements existing national surveillance data by providing detailed insights into adolescent nicotine use patterns, self-reported associated health impacts, and perceived parental awareness. These novel findings, particularly regarding high rates of nicotine use during nighttime awakenings and self-reported health complaints, offer valuable new perspectives that have been underexplored in previous research. By capturing these nuanced aspects of nicotine behaviour within a school setting, our study provides a stronger evidence base to inform the development of targeted and effective prevention and intervention strategies.

Several limitations of this study should be acknowledged. First, data were self-reported, which may introduce recall bias and social desirability bias, potentially affecting the accuracy of reported nicotine use and related behaviours. Second, participation bias cannot be excluded, as students who chose to participate might differ systematically from those who did not, possibly affecting generalizability. Third, the cross-sectional design limits causal inference regarding progression from experimentation to regular use. Fourth, although the overlap between the school-wide and targeted samples is expected to be minimal, it cannot be entirely ruled out, which may affect independence of observations. Finally, the relatively young average age of the sample influences the reported mean age of initiation, as age of initiation naturally depends on the age composition of the sample. A sample skewed toward younger adolescents will likely yield a lower mean age of initiation compared to a broader age range.

It is important to note that our study is not designed to provide definitive prevalence estimates on nicotine use, as the sample is unlikely to be fully representative of the entire Dutch adolescent population. The participating schools had relatively low Achterstandsscores, indicating a socioeconomically advantaged population. Furthermore, participation in the pilot project with *Vapen #jouwkeuze* was voluntary, which may reflect heightened awareness or concern regarding nicotine use, potentially due to higher prevalence in these schools.

The observed 28% past-year nicotine use prevalence in our school-wide survey likely underestimates the true prevalence within these school populations. This underestimation may be partly due to the exclusion of nicotine-using students recruited through targeted surveys, who represent a substantial subgroup. Additionally, the shorter questionnaires for non-users could have increased their response rates, thereby biasing prevalence estimates downward. The underrepresentation of older, final-year students, who were less available due to exam schedules, may also contribute to an underestimate of true prevalence. Consequently, the actual prevalence of nicotine use across the participating schools is expected to exceed 28%. Despite a two-year gap between datasets, the similarity between our school-wide prevalence and the 2023 national estimates (15) suggests that our sample reasonably approximates the broader adolescent population. However, caution is warranted when generalizing these findings nationally, given the non-random sampling of schools and students.

### Implications and future research

Nicotine use is widespread across diverse age groups, educational tracks, and genders, highlighting its status as a major public health concern. The typical initiation age marks a critical window for preventive interventions before and during early secondary education. There is a pressing need for comprehensive school-based prevention and cessation programmes, supported by strategies to enhance parental awareness and encourage family involvement. However, our data confirm that, to date, both schools and parents have been unable to effectively prevent or reduce nicotine addiction among youth. Given the availability of these highly addictive nicotine products for adolescents, experimentation during adolescence appears almost inevitable. In most cases, experimentation leads to regular use. Therefore, policymakers must take primary responsibility for limiting the tobacco industry’s influence and take actions to reduce youth access to nicotine products. Future research should investigate the effectiveness of school-based cessation interventions and identify pathways to prevent adolescent nicotine use on a national scale.

## Data Availability

All data produced in the present work are presented within the manuscript. Further details can be provided by the authors on reasonable request.

## LIST OF ABBREVIATIONS

CBS: Statistics Netherlands
HAVO: Senior General Secondary Education
VMBO: Pre-vocational Secondary Education
VWO: Pre-university Education
WMO: Dutch Medical Research with Human Subjects Act

## ACKNOWLEDGEMENTS

We sincerely thank the students and staff of the participating schools for their invaluable cooperation. Their involvement was crucial in enabling the successful completion of this research and in providing valuable insights into nicotine use among Dutch adolescents. We also express our gratitude to Hilda Teengs Gerritsen and Simone Mulders from the organising platform Vapen #JouwKeuze (www.vapenjouwkeuze.nl) for their ongoing collaboration and assistance.

## AUTHORS’ CONTRIBUTION

Daphne C.J. Raad, Danielle Cohen and David van Bodegom contributed to the study conception and design. Data collection and analysis were performed by Daphne C.J. Raad, Anne Marit Koome, and Yousef el Baser. The first draft of the manuscript was written by Daphne C.J. Raad and Danielle Cohen and all authors commented on previous versions of the manuscript. All authors read and approved the final manuscript.

## APPENDIX 1. Survey questionnaire

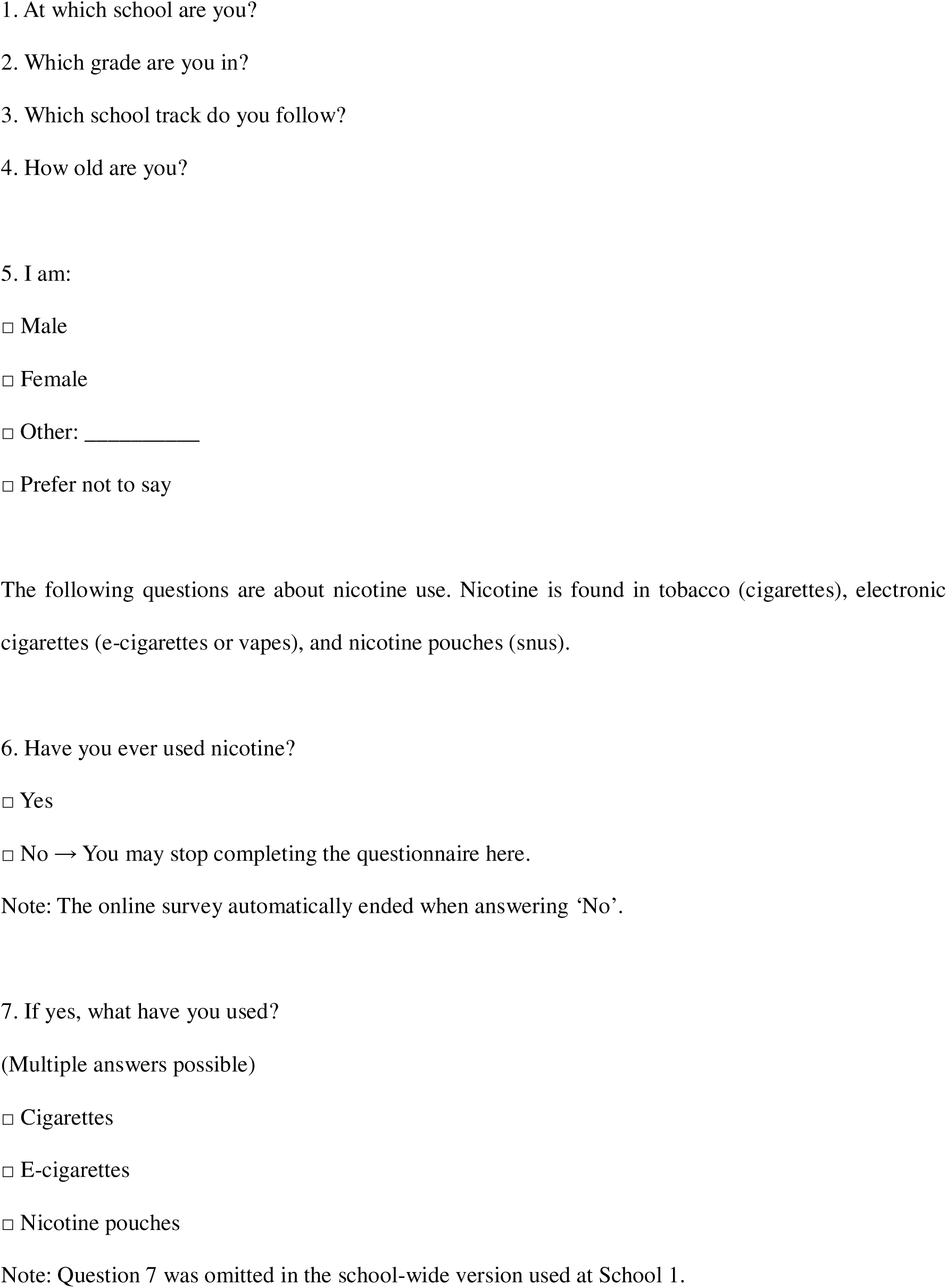
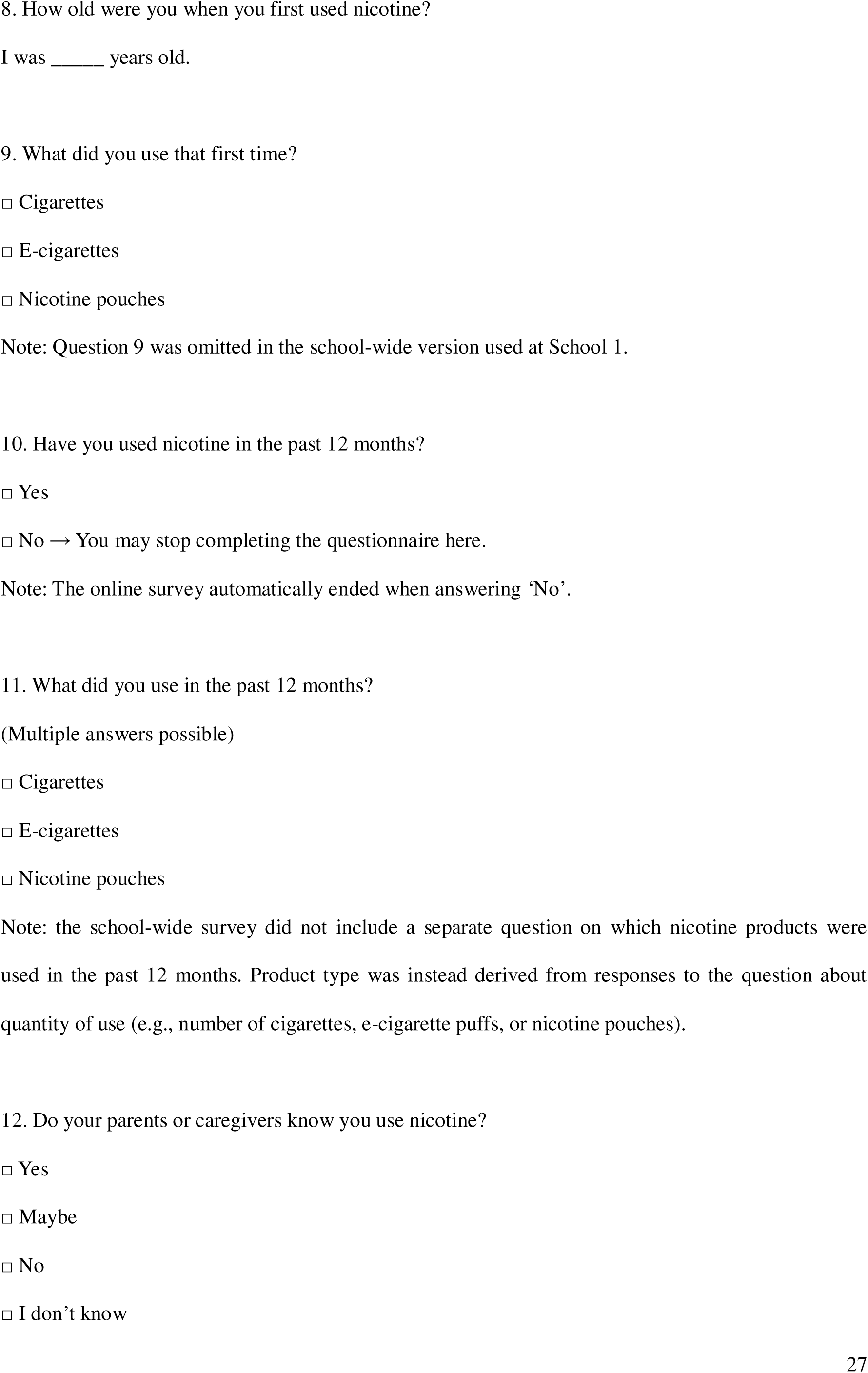
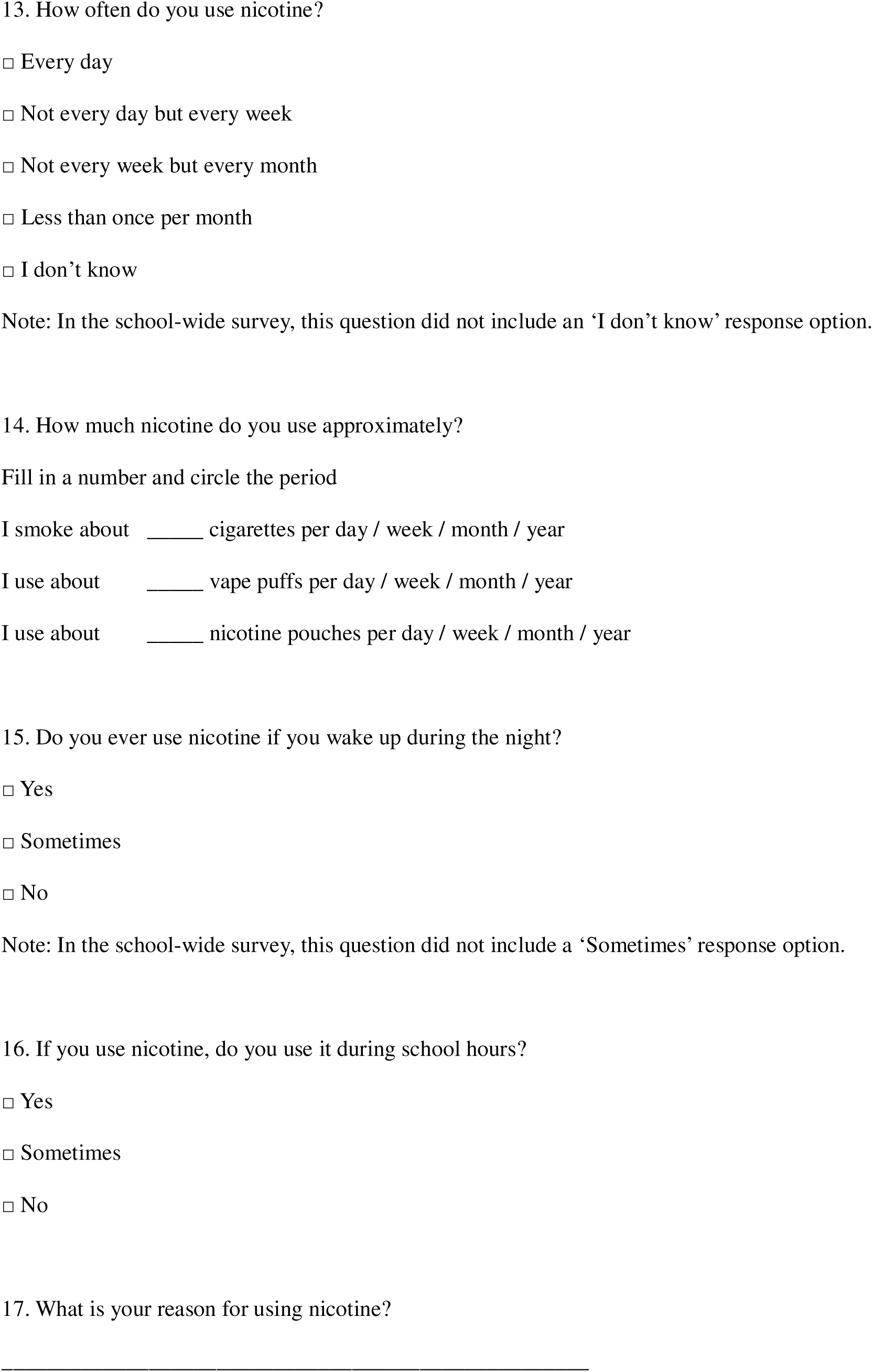
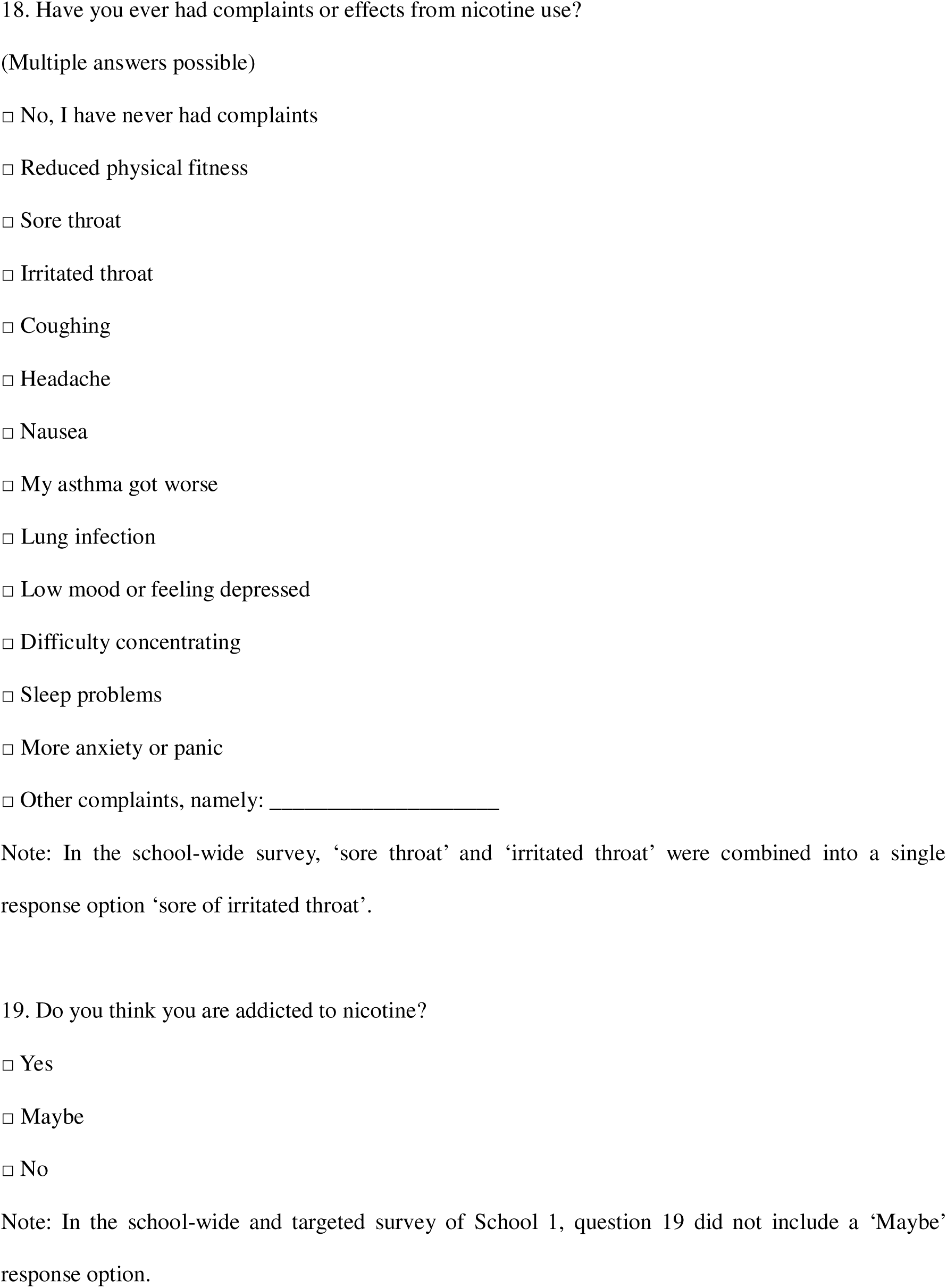
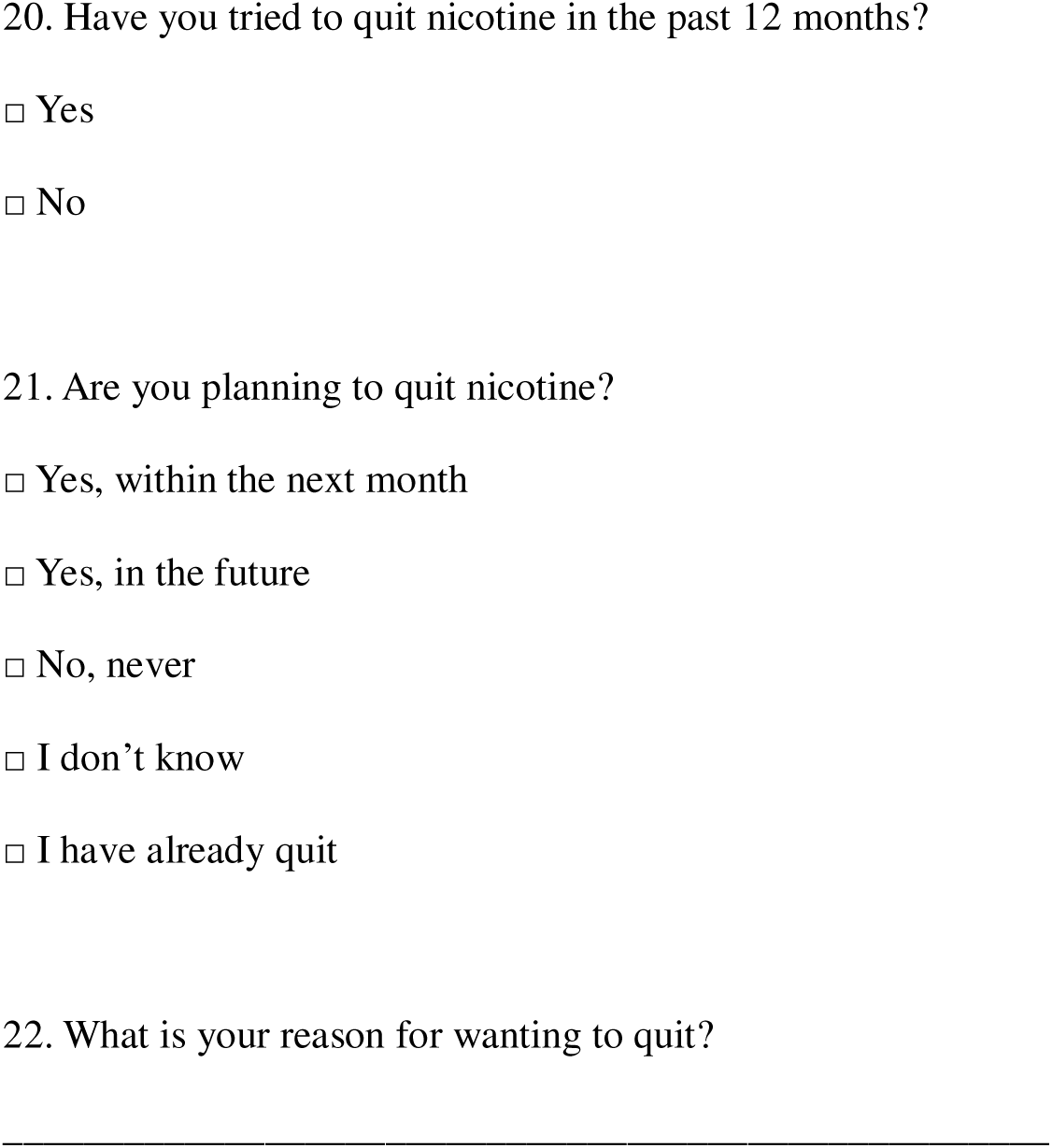

## REFERENCES

1. Boer M, van Dorsselaer S, de Looze M, de Roos S, Brons H, van den Eijnden R, et al. HBSC 2021: Gezondheid en welzijn van jongeren in Nederland. Utrecht 2022.

2. Rombouts MM, K.; van Dorsselaer, S.; Tuithof, M.; Monshouwer, K. Peilstationsonderzoek Scholieren 2023. Utrecht; 2023.

3. Jamal A, Gentzke A, Hu SS, Cullen KA, Apelberg BJ, Homa DM, et al. Tobacco Use Among Middle and High School Students - United States, 2011-2016. MMWR Morb Mortal Wkly Rep. 2017;66(23):597-603.

4. Huang J, Duan Z, Kwok J, Binns S, Vera LE, Kim Y, et al. Vaping versus JUULing: how the extraordinary growth and marketing of JUUL transformed the US retail e-cigarette market. Tob Control. 2019;28(2):146–51.

5. Guerra Castillo C, Hoeft KS, Couch ET, Urata J, Halpern-Felsher B, Chaffee BW. Adolescents’ Experiences and Perceptions of E-Cigarettes and Nicotine Addiction. Subst Use Misuse. 2024;59(13):1981–9.

6. Grana R, Benowitz N, Glantz SA. E-cigarettes: a scientific review. Circulation. 2014;129(19):1972–86.

7. Gordon T, Karey E, Rebuli ME, Escobar YH, Jaspers I, Chen LC. E-Cigarette Toxicology. Annu Rev Pharmacol Toxicol. 2022;62:301–22.

8. Martinelli T, Candel M, de Vries H, Talhout R, Knapen V, van Schayck CP, et al. Exploring the gateway hypothesis of e-cigarettes and tobacco: a prospective replication study among adolescents in the Netherlands and Flanders. Tob Control. 2023;32(2):170–8.

9. Miech R, Patrick ME, O’Malley PM, Johnston LD. E-cigarette use as a predictor of cigarette smoking: results from a 1-year follow-up of a national sample of 12th grade students. Tob Control. 2017;26(e2):e106–e11.

10. Soneji S, Barrington-Trimis JL, Wills TA, Leventhal AM, Unger JB, Gibson LA, et al. Association Between Initial Use of e-Cigarettes and Subsequent Cigarette Smoking Among Adolescents and Young Adults: A Systematic Review and Meta-analysis. JAMA Pediatr. 2017;171(8):788–97.

11. Barrington-Trimis JL, Urman R, Berhane K, Unger JB, Cruz TB, Pentz MA, et al. E-Cigarettes and Future Cigarette Use. Pediatrics. 2016;138(1).

12. Khouja JN, Suddell SF, Peters SE, Taylor AE, Munafo MR. Is e-cigarette use in non-smoking young adults associated with later smoking? A systematic review and meta-analysis. Tob Control. 2020;30(1):8–15.

13. Chan GCK, Stjepanovic D, Lim C, Sun T, Shanmuga Anandan A, Connor JP, et al. Gateway or common liability? A systematic review and meta-analysis of studies of adolescent e-cigarette use and future smoking initiation. Addiction. 2021;116(4):743–56.

14. Mongilio JM, Staff J, Seto CH, Maggs JL, Evans-Polce RJ. Risk of adolescent cigarette use in three UK birth cohorts before and after e-cigarettes. Tob Control. 2025.

15. Trimbos-instituut. Jongerenmonitor: Gebruik van tabaks- en nicotineproducten onder jongeren. Trimbos-instituut; 2023. Report No.: AF2112.

16. Stolerman IP, Jarvis MJ. The scientific case that nicotine is addictive. Psychopharmacology (Berl). 1995;117(1):2–10; discussion 4-20.

17. Becker TD, Arnold MK, Ro V, Martin L, Rice TR. Systematic Review of Electronic Cigarette Use (Vaping) and Mental Health Comorbidity Among Adolescents and Young Adults. Nicotine Tob Res. 2021;23(3):415–25.

18. Mukerjee R, Hirschtick JL, Arciniega LZ, Xie Y, Barnes GD, Arenberg DA, et al. ENDS, Cigarettes, and Respiratory Illness: Longitudinal Associations Among U.S. Youth. Am J Prev Med. 2024;66(5):789–96.

19. Palamidas A, Tsikrika S, Katsaounou PA, Vakali S, Gennimata SA, Kaltsakas G, et al. Acute effects of short term use of ecigarettes on Airways Physiology and Respiratory Symptoms in Smokers with and without Airway Obstructive Diseases and in Healthy non smokers. Tob Prev Cessat. 2017;3:5.

20. England LJ, Bunnell RE, Pechacek TF, Tong VT, McAfee TA. Nicotine and the Developing Human: A Neglected Element in the Electronic Cigarette Debate. Am J Prev Med. 2015;49(2):286–93.

21. Do EK, Tulsiani S, Koris K, Minter T, Hair EC. Depression, anxiety, stress, and current e-cigarette use: Results from the Truth Longitudinal Cohort of youth and young adults (2022-2023). J Affect Disord. 2024;365:628–33.

22. Williams MB, K. N.; Talbot, P. Analysis of the elements and metals in multiple generations of electronic cigarette atomizers. Environmental Research. 2019;175:156–66.

23. Gorfinkel L, Hasin D, Miech R, Keyes KM. The Link Between Depressive Symptoms and Vaping Nicotine in U.S. Adolescents, 2017-2019. J Adolesc Health. 2022;70(1):133-9.

24. Bandiera FC, Loukas A, Wilkinson AV, Perry CL. Associations between tobacco and nicotine product use and depressive symptoms among college students in Texas. Addict Behav. 2016;63:19–22.

25. #jouwkeuze V. Vapen #jouwkeuze 2025 [Available from: https://vapenjouwkeuze.nl/.

26. Borm FJ, Cohen S, Milani GP, de Winter P, Cohen D. I will prevent disease whenever I can, for prevention is preferable to cure: doctors’ role in the vaping epidemic. Eur J Pediatr. 2024;183(6):2517–20.

27. Branstetter SA, Krebs NM, Muscat JE. Nighttime Waking to Smoke, Stress, and Nicotine Addiction. Behav Sleep Med. 2022;20(6):706–15.

28. Houterman K, van Roosmalen M, van Hijfte R. Deze tieners lagen in ziekenhuis na vapen: ‘Extreem benauwd, bloed ophoesten en overgeven’ RTL Nieuws 2025 [Available from: https://www.rtl.nl/nieuws/binnenland/artikel/5506933/ook-tim-gabriel-fenna-gijs-en-jayden-lagen-ziekenhuis-na-vapen.

29. Belok SH, Parikh R, Bernardo J, Kathuria H. E-cigarette, or vaping, product use-associated lung injury: a review. Pneumonia (Nathan). 2020;12:12.

30. Vroemen M, Tromp J, Boesschen Hospers J. De illegale vapehandel op schoolpleinen neemt toe, maar makkelijk verdienen is moeilijk te stoppen. De Volkskrant. 2025 22 March 2025.

31. Rifat MA, Orsini N, Qazi B, Galanti MR. Smoking Prevention and Cessation Programs for Children and Adolescents Focusing on Parental Involvement: A Systematic Review and Meta-Analysis. J Adolesc Health. 2025;76(4):532–41.

